# The Burden of Long COVID: Re-Infection, Symptoms, and Barriers to Care in the Arizona CoVHORT Study

**DOI:** 10.64898/2026.08.18.26360693

**Authors:** Kristen Pogreba-Brown, Caitlyn McFadden, Kelly M. Heslin, Dametreea L. Carr, Laura P. Falk, Collin Catalfamo, Kacey Ernst, Leslie V Farland, Felinia Cordova-Marks, Xiaoxiao Sun, Leila Barraza, Erika Austhof, CoVHORT Study Group

**Author notes:** **Corresponding Author Information** Kristen Pogreba-Brown, PhD MPH 1295 N. Martin Avenue, PO Box 245211 Tucson, AZ, USA 85724-5211.

## Abstract

Long COVID (LC) impacts quality of life, daily functioning, and healthcare utilization. Understanding the onset and duration of symptoms, characteristics of those at higher risk, and the barriers and facilitators for healthcare access and therapeutics are key to addressing this growing disease burden. In 2024 the Arizona CoVHORT, an ongoing 6-year longitudinal study, distributed a cross-sectional LC survey to gain additional in-depth information. Of 1,543 participants, 700 reported LC symptoms lasting 2-49 months. Following their first infection, LC+ participants had a 21% higher risk for a second infection and were 3.2 times more likely to report LC symptoms after that second infection compared to LC-participants. Significant factors associated with LC included female sex (OR=2.3), Hispanic ethnicity (OR=1.5), BMI>34.5 (OR= 1.7) and >2 infections (OR=3.2), while vaccination prior to first infection decreased the odds of reporting LC by 51% (R=0.49). Qualitative analyses detailed significant barriers to care and encounters with providers who lacked knowledge to test for or treat LC symptoms. With an estimated 400 million people impacted globally by LC, it is critical to gain in-depth information from patients to improve both access and quality of care, improve messaging, and target mitigation strategies to decrease the burden over time.

## INTRODUCTION

Prolonged symptoms associated with Long COVID (LC) can impair multiple organ systems and commonly include fatigue, cognitive impairment, joint pain, poor mental health, and gastrointestinal symptoms, among others.[1] Current studies on LC suggest female sex, greater number of comorbidities, older age, and disease severity during acute phase of infection are associated with increased risk of developing the condition. [1–3]

Prevalence estimates of LC vary substantially based on population studied, timing of the study, and dynamic nature of the pandemic and virus. A recent meta-analysis of 429 studies found the prevalence to be 30% in the United States (US) and 36% in a global pooled estimate.[4] Two large cross-sectional studies in 2022 estimated 11-14% of the US population has ever experienced LC and approximately 15-17 million (7%) people are still reporting symptoms.[5,6] Worldwide, the cumulative global incidence is 400 million people, adding to the growing burden of global chronic disease.[7] Economically, the burden is estimated at over $1 trillion annually with annual lost wages in the US to be $170 billion.[6]

While LC is recognized within the medical community, there are still few resources for patients looking to mitigate the impacts of their symptoms. International publications on LC patient experiences have detailed various barriers to accessing quality healthcare including limited awareness of potential LC resources and medical expertise, difficulty navigating healthcare systems, poor care continuity, and experiencing stigma.[8–10] Investigations into US-based patient experiences, though scarce, have reported similar barriers in addition to high cost of care.[11] Despite these findings, granular insights into the lived experience of LC in the US remain limited.

This paper describes demographics, vaccination history, infection count, and onset of symptoms among people with LC (LC+) compared to those infected, but who reported no post-acute symptoms (LC-). Among those with LC+, we examined healthcare utilization, provider interactions, and self-reported symptom management. These results can be used by clinicians, patient advocates, public health practitioners and others to determine the challenges and needs of this growing population.

## MATERIALS and METHODS

### Survey Design and Distribution

To better understand the acute and long-term impacts of COVID-19, the Arizona CoVHORT cohort study, was initiated in May 2020, as described previously.[12] Briefly, participants were recruited through multiple channels and eligibility was open to all state residents 18+ regardless of COVID-19 history.

Routine quarterly surveys were administered through REDCap (hosted at University of Arizona)[13,14] to collect demographics, chronic health conditions, infection status, vaccination history, symptoms (acute and post-acute), mental health, and household information. To further assess the secondary impacts of LC, including day-to-day activities, financial burden, ability to work and more, a cross-sectional ‘Living with Long COVID’ survey was emailed to participants between January 27-May 12, 2024. We included 4,794 of 9,090 (52.7%) CoVHORT participants who had indicated a previous SARS-CoV-2 infection, 518 (10.8%) of whom indicated they were clinically or self-diagnosed with LC on a previous quarterly CoVHORT survey.

The complete survey is available on GitHub (github.com/CoVHORT-Study/LivingWithLongCovid) with pertinent questions summarized below. Screening questions identified people who had a confirmed or suspect SARS-CoV-2 infection since January 2020. For each infection (up to six), participants were asked to recall the month and year of symptom onset/positive test, vaccination status at time of infection, and symptoms lasting 30 days or more after each infection. Symptoms/conditions were broken down into three categories:

1. ONGOING (*any symptom that started during your COVID illness and never got better*),
2. WORSENING (*any symptom you may have had before you got COVID that got worse after your COVID illness*) or
3. NEW symptoms (*any symptom that started after you felt like you had recovered from your initial COVID illness*)

The categories were applied across 48 commonly reported LC symptoms in a multi-select checkbox format, including an open text ‘other’ field. Participants reporting no symptoms at 30 days post-infection could skip to the closing sections. Participant demographics (age, sex, race, ethnicity) were collected at baseline.

### Determination of Long COVID and Vaccination Status

Participants were classified as LC+ if they reported new, ongoing or worsening symptoms a month or more after infection. Symptom duration was determined by resolution date and calculated as the time between infection onset and symptom resolution. Those with ongoing symptoms were censored using survey completion dates. Participants who recovered within one month or reported no new, ongoing, or worsening symptoms were classified as LC-. LC status following each infection was determined and combined to assess for LC at any point since 2020. Vaccination status two weeks or more prior to each infection was recorded, but only vaccination prior to the first infection was included in analyses.

### Statistical Analyses

To describe our sample, participant demographics, number of COVID infections, and vaccination history were stratified by LC status. Categorical variables were summarized by frequencies and proportions, and continuous data as means (SD). Demographic covariates included sex and gender (female, male, transgender), age (continuous), race (American Indian/Alaska Native, Asian, Black or African American, white, mixed race), ethnicity (non-Hispanic, Hispanic) and body mass index (BMI; <35kg/m^2^, ≥35kg/m^2^).

To describe and characterize LC symptoms, average symptom duration and range in months were estimated for LC+ participants, stratified by infection number and recovery status. First infections were prioritized here given the longest follow up time from infection to survey date, allowing for the most complete picture of symptom data. The frequency and proportion of participants reporting various healthcare interactions is described. We also conducted a multivariable logistic regression analysis to further investigate the associations between key demographic factors, vaccination history at first infection, and number of infections and long COVID status. Race was excluded due to small sample size among specific groups, and the number of infections was collapsed into 2 levels (1-2 and 3-6) to facilitate model stability.

### Qualitative Analyses

To support quantitative findings, thematic analysis of three free-response questions was conducted to identify quotes representative of participant experiences:

1. Of all the things you tried to help resolve your symptoms, what do you think helped the most?
2. What barriers did you experience when seeking care or after you received care?
3. What was your experience like when seeking care?

Analyses followed recommended thematic analysis steps[15,16] and were performed by two independent coders (DC and KH) to enhance trustworthiness and rigor.

Coding Process:

1. Familiarization with and organization of data
2. Inductive coding of responses into keywords, concepts, and themes
3. Discussion and comparison of independent codes, resolution of coding differences, and discussion of biases to reach mutual agreement and develop consensus themes
4. Identification of seminal quotes that support each theme describing LC experiences All data analyses were conducted using Stata 18 (College Station, TX), RStudio v2025.05.0+496 (Boston, MA), and Microsoft Excel v.2508. This study was approved by the University of Arizona Internal Review Board (#2003521636).

## RESULTS

### Demographics

From January to June 2024, 1,709 CoVHORT participants completed the ’Living with Long Covid’ survey with 1,543 who had a previous COVID infection and provided enough illness and symptom data to determine their long COVID status. Of those, 843 (54.7%) reported no continuing symptoms 30 days or longer after their acute infection (LC-) and 700 (45.3%) reported at least one or more longer-term symptom (LC+) (Figure 1). Of the 700 LC+ participants, 568 (81.0%) reported symptoms lasting 3 months or longer and 164 (23.0%) reported an LC diagnosis by a clinician.

The mean age was 51.5 years for LC+ and 53.2 years for LC-participants. Women made up a larger proportion of the LC+ group (79.0%) compared to the overall distribution of respondents (70.1%) (Table 1). A total of 162 (10.9%) participants were Hispanic, with 89 (13.4%) in LC+ and 73 (8.8%) in LC-. Participants with a BMI ≥35kg/m^2^ comprised a larger proportion of the LC+ group (19.5%) compared to the LC-group (9.5%).

**Table 1.** Demographics, number of infections, and vaccination status at first infection among CoVHORT participants by long COVID status (n = 1,543)

| Characteristic | LC+<br>N (%) | LC-<br>N (%) | Total<br>N (%) |
| --- | --- | --- | --- |
| **Participants** | 700 (45.4) | 843 (54.6) | 1,543 (100.0) |
| **Age** (mean, SD) | 51.5 (14.42) | 53.2 (14.84) | 52.4 (14.67) |
| Female | 527 (79.0) | 523 (62.9) | 1,051 (70.1) |
| Male | 130 (19.5) | 299 (35.9) | 429 (28.6) |
| Non-Binary /Transgender Female<br>/Transgender Male /Prefer not to answer* | 10 (1.4) | 10 (1.2) | 20 (1.3) |
| American Indian/Alaska Native | <10 | <5 | <10 |
| Asian | 10 (1.5) | 26 (3.1) | 36 (2.4) |
|  | <10 (~1) | 10 (1.2) | 17 (1.1) |
| Black of African American |  |  |  |
| White | 604 (91.1) | 762 (91.6) | 1,367 (91.4) |
| Mixed Race | 23 (3.5) | 23 (2.8) | 46 (3.1) |
| Prefer not to answer | 11 (1.7) | 10 (1.2) | 21 (1.4) |
| Non-Hispanic | 569 (86.0) | 747 (90.4) | 1,317 (88.4) |
| Hispanic | 89 (13.4) | 73 (8.8) | 162 (10.9) |
| Prefer not to answer | <5 | <10 | 10 (0.7) |
| **BMI** |  |  |  |
| <35 | 564 (80.6) | 763 (90.5) | 1327 (86.0) |
| ≥ 35.0 | 136 (19.4) | 80 (9.5) | 215 (13.9) |
| **Vaccinated prior to 1<sup>st</sup> infection** |  |  |  |
| Yes | 391 (55.9) | 615 (73.0) | 1,006 (65.2) |
| No | 302 (43.1) | 220 (26.1) | 522 (33.8) |
| **Number of SARS-CoV-2 infections** |  |  |  |
| 1 | 322 (46.0) | 569 (67.5) | 891 (57.7) |
| 2 | 269 (38.4) | 233 (27.6) | 502 (32.5) |
| 3 | 85 (12.1) | 36 (4.3) | 121 (7.8) |
| 4 | 14 (0.2) | 4 (0.5) | 18 (1.2) |
| 5 | 6 (0.8) | 1 (0.1) | 7 (0.5) |
| 6 | 4 (0.6) | 0 (0) | 4 (0.3) |

\*Collapsed due to small sample sizes\*\*LC+ defined as reporting symptoms for more than one month after each infection. Not mutually exclusive; people may report extended symptoms after multiple infections. Percentage of LC+ developed for each numbered infection based on participants with complete data on infection dates.

### Multiple Infections & History of Vaccination

Incidence and Risk of LC by Infection: Amongst 1,543 participants, 2,389 infections were reported from January 2020 to May 2024. For these analyses, we defined incident LC as the first report of LC symptoms meeting the inclusion criteria. Prevalent LC was defined as any subsequent LC instances after the incident one, regardless of reported recovery status between instances. The incidence of LC was 35.6% (n=550) among people following a first infection (n=1,543) (Figure 2). Following a second infection (n=652), the incidence of LC was 10.1% (n=66) and the prevalence was 21.9% (n=143). For a third infection, the incidence and prevalence of LC+ was 4.7% (n=7) and 28.0% (n=42) respectively (Table S1). For people who developed LC following their first infection, the risk of reporting LC symptoms after a second infection was 3.2 times higher compared to people without a history of long COVID (RR=3.23; 95% CI 2.52-4.13).

Risk of Re-Infection by LC History: Most participants reported 1-2 infections (84.4% (LC+) and 95.1% (LC-)). For people who had 3-6 infections, 4.9% reported never having LC compared to 14.6% LC+ (Table 1). This relationship was also clear in the univariate modeling where each subsequent infection increased the odds of reporting a history of long COVID by approximately 2-fold (Table 2). Data from this survey also showed the risk of a second infection was 21% higher for people who had developed LC after their first infection compared to people who had not (RR=1.21; 95% CI 1.08-1.36).

**Table 2.** Factors associated with the odds of reporting prevalent Long COVID following ≥1 SARS-CoV-2 Infection – Univariate and multivariable logistic regression models (n=1,439)

| Dependent Variables | Univariate<sup>1</sup> | Multivariable<sup>2</sup> |
| --- | --- | --- |
| Age | 0.99 (0.99-1.0) | 0.99 (0.99-1.0) |
| Female | 2.32 (1.82-2.94) | 2.33 (1.81-3.01) |
| Hispanic | 1.60 (1.15-2.22) | 1.49 (1.04-2.14) |
| Vaccinated prior to first infection | 0.46 (0.37-0.57) | 0.49 (0.39-0.62) |
| BMI (binary with <34.5 as ref) | 2.30 (1.71-3.10) | 1.72 (1.21-2.43) |
| # Infections ($\leq 2$ ref vs 3+)* | 3.61 (2.48-5.25) | 3.18 (2.08-4.84) |
| # infections (1=ref) |  |  |
| 2 | 2.04 (1.63-2.55) |  |
| 3 | 4.17 (2.76-6.30) |  |
| 4 | 6.18 (2.02-18.95) |  |
| 5 | 10.60 (1.27- |  |
| 6 | 88.46) |  |
|  | NA |  |

<sup>1</sup>Univariate modeling for reporting long COVID after $\geq 1$ infection (dependent) each potential predictor (independent variable)
<sup>2</sup>Multivariable model for reporting long COVID after $\geq 1$ infection (dependent) with all potential predictors included in the model
\*Number of infections included in the model as a binary variable with 1-2 infections (ref) compared to 3-6 infections. This was used in place of number of infections in the multivariable model for ease of interpretation, though the results by infection number were similar in the multivariate as the univariate models (results not shown).

When asked about being reinfected after already developing LC, 56% reported their LC symptoms were largely unchanged, although 30% reported their symptoms got worse, 3% reported their symptoms got better, and 10.6% reported their symptoms changed.

Vaccination History: Among all the participants, 65.2% reported being vaccinated 2 or more weeks before their first infection. Amongst LC+ participants, 55.9% were vaccinated at least 2 weeks prior to their first infection compared to 73.0% of LC-participants (Table1). Vaccination was also found to be protective against LC in both the univariate and multivariable models – see below (Table 2).

### Length of Symptoms and Resolution

On average, 63% (n=346) of LC+ participants reported active symptoms 29 months after their first infection (range 2-49 months). Among those with LC symptom resolution following their first infection, the mean length of symptoms was 7.2 months (range 2-42 months). Subsequent infections resolved in slightly less time, but were also represented by much smaller groups (n=48 and n=7 respectively) (FIGURE 2 with additional data in Table S1).

### Risk Factors for Long COVID Modeling

Table 2 shows the results of a logistic model comparing LC+ to LC-participants. In the univariate, unadjusted model, there was a clear dose-response relationship with number of infections and reporting of LC increasing the odds of reporting LC by approximately 2 times with each subsequent infection. After multivariable adjustment, female sex (OR=2.33; 95% CI 1.81-3.01) and Hispanic ethnicity (OR=1.49; 95% CI 1.04-2.14) were both associated with increased odds of reporting LC, while age was not (OR=1.0; 95% CI 0.99-1.00). Vaccination prior to first infection decreased the odds of reporting LC by 51% (OR=0.49; 95% CI 0.39-0.62) after adjusting for other variables. Compared to participants reporting one or two infections, having three to six infections increased the odds of reporting LC three-fold (OR=3.18; 95% CI 2.08-4.84) after adjusting for other variables.

### Long COVID Symptoms & Symptom Relief

The most commonly reported symptoms among LC+ participants were fatigue (72.4%), brain fog (57.4%), aches and pains (48.3%), shortness of breath (46.0%), and cough (43.2%). Regarding onset type of LC symptoms, the majority of participants (84%) reported at least one ongoing symptom, compared to 40% who reported a new symptom after their acute phase, and 34% with pre-existing symptoms that became worse 1+ months after their infection. Participants most commonly reported 1-5 LC symptoms (n = 260), with proportions declining as symptom count increased, except for a moderate uptick among those reporting 26+ symptoms (n = 63) (Figure 3).

Regarding symptom relief, conventional medicine was the most commonly reported free-text response. Additional activities included sleep and rest; diet, nutrition, exercise and active lifestyle; home remedies/alternative methods; and patience with recovery time. Quotes representative of common actions taken include:

*“Eating right, exercise and taking supplements regularly like vitamin D, Calcium and Minerals”, “Acupuncture and massage,”*

*“breathing exercises,”*

*“…staying home and resting…”* and

*“Just letting the symptoms resolve over time…”*

When asked if anything specific contributed to feeling better and what helped the most to resolve symptoms, time was reported more often by people who had fully recovered (n=139) compared to those who had not (n=80). Helpful activities reported by those who had not fully recovered (n=310) included getting enough sleep (n=180), gentle exercise (n=167), and altering nutrition or eating habits (n=104). Notably, in comparison to those who had fully recovered, the not fully recovered group also reported taking a larger number of dietary supplements (n=83 vs. 26), physical therapy and rehabilitation (n=49 vs. 8), other therapies (n=54 vs. 10), and other treatments (n=67 vs. 26).

### Healthcare Use and Barriers

For participants who sought care from a healthcare provider (n=563), the majority saw a provider ≥2 times to manage symptoms (25.3%). In addition to being told by a healthcare provider that they had LC (30.6%), participants reported being diagnosed with other related conditions including sleep disturbances, postural orthostatic tachycardia syndrome (POTS), irritable bowel syndrome (IBS), Myalgic Encephalomyelitis/Chronic Fatigue Syndrome (ME/CFS), reactivation of Epstein-Barr Virus, and dyspnea or fibrosis, among 82 other diagnoses (Table S2). Specialists most commonly seen by LC participants included cardiology (n=175), neurology (n=99), internal medicine (n=86), physical therapy (n=81), gastroenterology (n=80), and psychology (n=80). To seek care, 72 (12.8%) participants traveled ≥1 hours (mean 3hr, range 1-30) for appointments with primary care (n=14), specialists (n=70), or other providers (n=7).

Participants experienced significant barriers when seeking and receiving care, including delays securing appointments, challenges with geographic proximity to appointment locations, encounters with providers who lacked knowledge to support or treat LC symptoms, anxiety, and their LC not being taken seriously by providers (Figure 4). Participants also reported limited testing and treatment options when they did obtain appointments. Specifically, one participant noted:

*“[M]y dr doesn’t know how to diagnose or help me.”*

Other representative quotes include:

*“I had to beg for basic care. Some doctors believed me and some did not. I have so much anxiety about appointments and got so little real care I stopped going. I have basically given up;”*

*“My experience with seeking care has been extremely poor. Medical providers are not taking my LC seriously.”*.

While many people reported difficulties, several others noted the importance of having a supportive provider to help navigate their care and ongoing symptoms. When asked what helped the most in treating their symptoms, many acknowledged medical professionals who worked with them to find answers. Representative quotes include:

*“Doctors who actively went above and beyond and didn’t give up and kept doing tests.”*

*“Knowing that my primary care provider is there for me if and when I need him. He not only provides physical [c]are but relieves any anxieties I may be experiencing.”*

*“The eagerness of one practitioner to help in any way possible I’m so grateful I found my practitioner”*

*“Knowing my doctor is taking me seriously and being consistent with all of my maintenance meds”*

## DISCUSSION

We administered a cross-sectional survey within the larger AZ CoVHORT cohort study to gain a better understanding of participants’ LC experiences and identify risk factors compared to those who were infected but did not develop LC. Overall, we found that vaccination prior to the first infection was protective. Other factors reported more often in the LC+ group included female sex, Hispanic ethnicity, BMI over 34.9kg/m^2^ and reporting multiple infections since 2020. These results are consistent with several studies and systematic reviews that have found an increased risk for LC among female sex specific co-morbidities, severe acute infection, being unvaccinated prior to infection, and multiple infections.[2,17–19] Regarding ethnicity, our results found a 49% higher odds of LC among Hispanic participants which has not been widely reported among self-reported surveys.[20] This may be due in part to limited awareness of LC and its symptoms.[21]

Prolonged symptoms reported most frequently among those with LC included fatigue, brain fog, aches and pains, and shortness of breath, with those who recovered having an average time to symptom resolution of 7.2 months (range 2-42 months) following the first infection. Healthcare barriers with seeking and receiving care were consistently reported by participants, underscoring the need for additional resources to reduce this disease burden as these infections continue to arise.

### Number of Infections

Infection increased the odds of LC by approximately twofold with each additional infection, consistent with findings from previous studies.[19,22,23] However, given the nature of these analyses, this relationship may operate in multiple ways, either by increasing reinfection susceptibility among those already LC+, which our results would imply, or by repeated infections over time increasing the risk of developing LC; or a combination of the two, which is biologically plausible. Evidence suggests that some people with LC exhibit hallmarks of immune dysregulation,[24–26] while multiple infections may impair viral clearance, resulting in ongoing viral persistence - another leading hypothesis for LC.[27,28] This dose-response relationship should be considered in future LC studies to better clarify possible drivers of risk and inform mitigation strategies. Within our sample, 79% of participants developed long COVID after their first infection, suggesting the higher odds of multiple infections may reflect increased susceptibility, potentially due to reduced immune function associated with LC. This is also supported by group comparison, where 16% of the LC+ group reported 3 or more infections compared to 4.9% of the LC-group.

### Vaccination History

Vaccination at least two weeks prior to first SARS-CoV-2 infection was less common among LC+ participants than LC- (55.9% vs 73.0%) and reduced the odds of developing LC by 51% after multivariable adjustment. This protective effect is consistent with many other studies worldwide,[2,22,29] including a large systematic review and meta-analysis reporting similar effect estimates (0.60 and 0.64 for one and two doses, respectively) when compared to people with LC who were not vaccinated.[30] Our results of 20% of people reporting improved LC symptoms following a SARS-CoV-2 vaccination is also consistent with the 20.3% reported within the same review.[30]

### Symptoms

Fatigue (72%) and brain fog (57%) were the most consistently reported symptoms amongst people with LC, followed by body aches, shortness of breath, cough, insomnia, and anxiety. Notably, these symptom prevalences were observed in a primarily non-clinical cohort, historically underrepresented in LC literature.[31,32] Only 2.5% of CoVHORT participants were hospitalized during acute illness and 21% sought medical care, proportions lower than those reported in similar studies.[33,34] These findings suggest that the burden of LC extends broadly across the severity spectrum and may better represent patients who do not seek care during their acute illness.

Our survey also distinguished symptom onset as new, ongoing or became worse a month or more after infection – a level of granularity underexplored in LC research. Among all 48 symptoms queried, ongoing symptoms were notably more prevalent than new or worsening symptoms. These findings suggest that LC is most often a continuation of symptoms that began during the acute infection, rather than symptoms that develop later or an exacerbation of pre-existing conditions, both of which have the potential for misclassification bias of LC if they are not related to the infection. This is consistent with our prior work showing that while people with pre-existing chronic conditions were at higher risk of developing LC, this is not simply due to an exacerbation of pre-existing symptoms.[3]

### Recovery

Estimating LC recovery time is extremely difficult outside of a longitudinal study design; this survey addressed this gap by allowing participants to provide a recovery date (month/year) for each infection listed. Recovery took longest after the first infection. On average, recovery time was 7.2 months; however, among those who had not yet recovered, symptoms persisted for approximately 29 months, indicating a substantial chronic burden. Notably, recovery time for those that reported ≥3 infections, may be a reflection of a shorter period of time from infection to survey completion rather than a faster resolution of symptoms. Future studies with longer follow-up periods will be able to address this more thoroughly.

### Activities of Daily Living & Healthcare Use

LC profoundly affects functional status. One in five participants reported their LC symptoms reduced their ability to carry out daily activities, with almost 70% reporting at least some impact. Reduced levels of participation in daily activities impact physical and mental well-being, social engagement, and employment, resulting in an overall reduction in quality of life.[35,36] The majority of participants who sought care to manage their LC symptoms saw providers multiple times. Concurrent diagnoses of other conditions related to immune function (e.g. IBS), chronic conditions (e.g. POTS, dyspnea) and a myriad of other comorbidities were also reported by one-third of LC+ participants, consistent with similar studies.[8,37] While seeking and receiving care from providers was a significant barrier to participants, experiences ranged from complete dismissal to sincere appreciation for the support and care received. These results are consistent with other qualitative LC studies.[38] As trusted sources of health information, it is imperative that healthcare providers be cognizant of their critical role in providing patient support and its impact on patient quality of life.[37]

### Strengths

This study has several strengths. First, as part of an existing COVID-19 study, we were able to focus this survey on more in-depth questions specifically related to LC, allowing us to gather data on potential LC risk factors from both LC cases and controls. Next, because our study primarily includes participants who did not seek medical care for their acute infection, these data align more closely with the broader population infected with SARS-CoV-2 and those who developed LC compared with LC studies based on hospitalized or EHR-based cohorts. Further, although over 90% of CoVHORT participants reported being vaccinated to-date, our study began in mid-2020, almost a year before the vaccine was widely available. Many participants therefore reported infections in 2020 and 2021, providing a large enough sample to determine the effect of vaccination status prior to infection. Finally, qualitative data analysis of open text fields provides deeper insight into patient impacts and may inform future clinical and public health approaches.

### Limitations

We acknowledge that selection bias may influence participation in both the CoVHORT study and this survey. However, the roughly equal proportions of LC+ and LC-participants allowed us to assess LC status and risk factors. As our sample size was relatively small, further research is needed to better understand how demographics and socioeconomic drivers, like access to healthcare, influence the risk of LC development and care. Regarding the questions on healthcare utilization and impacts on health and daily life, we did not include LC-because the questions were designed to address the effects of LC. Our core study assesses general health and well-being in a way that allows for these comparisons but was beyond the scope of this paper.

Given the survey’s timing, LC was defined according to CDC and NIH criteria in use at the time – symptoms lasting 28 days or longer after infection[39] - rather than the current National Academies definition of 90 days or longer of symptoms.[40] Because this definition was provided to participants, all analyses used the 28+ day definition, however a sensitivity analyses using 90+ days to determine LC status yielded similar results with a smaller sample size. This is useful for studies that were in progress before or during the development of the updated definition whereby the elevated risk factors, protective mechanisms, populations at risk and symptoms do not vary widely from earlier definitions.

### Conclusions

The goal of this cross-sectional survey within the Arizona CoVHORT Study was to gain more in-depth knowledge of the risks, symptoms and impacts to healthcare and day-to-day life, and functioning among our participants with LC. LC has been a complicated set of conditions to untangle, particularly without the aid of a clear diagnostic test and an extremely encompassing case definition. This study can help clinicians better understand the risks, experiences and barriers faced by people with LC to ultimately support patients in reducing the severity and burden of symptoms.

## STATEMENTS

### Data Availability Statement

A limited dataset and code for analyses for this study will be available 6-months following publication at github.com/CoVHORT-Study/LivingWithLongCovid.

Because many of these analyses depend on infection dates which are protected health data, these data can be available upon request.

## Data Availability

A limited dataset and code for analyses for this study will be available 6-months following publication at github.com/CoVHORT-Study/LivingWithLongCovid.
Because many of these analyses depend on infection dates which are protected health data, these data can be available upon request.

---

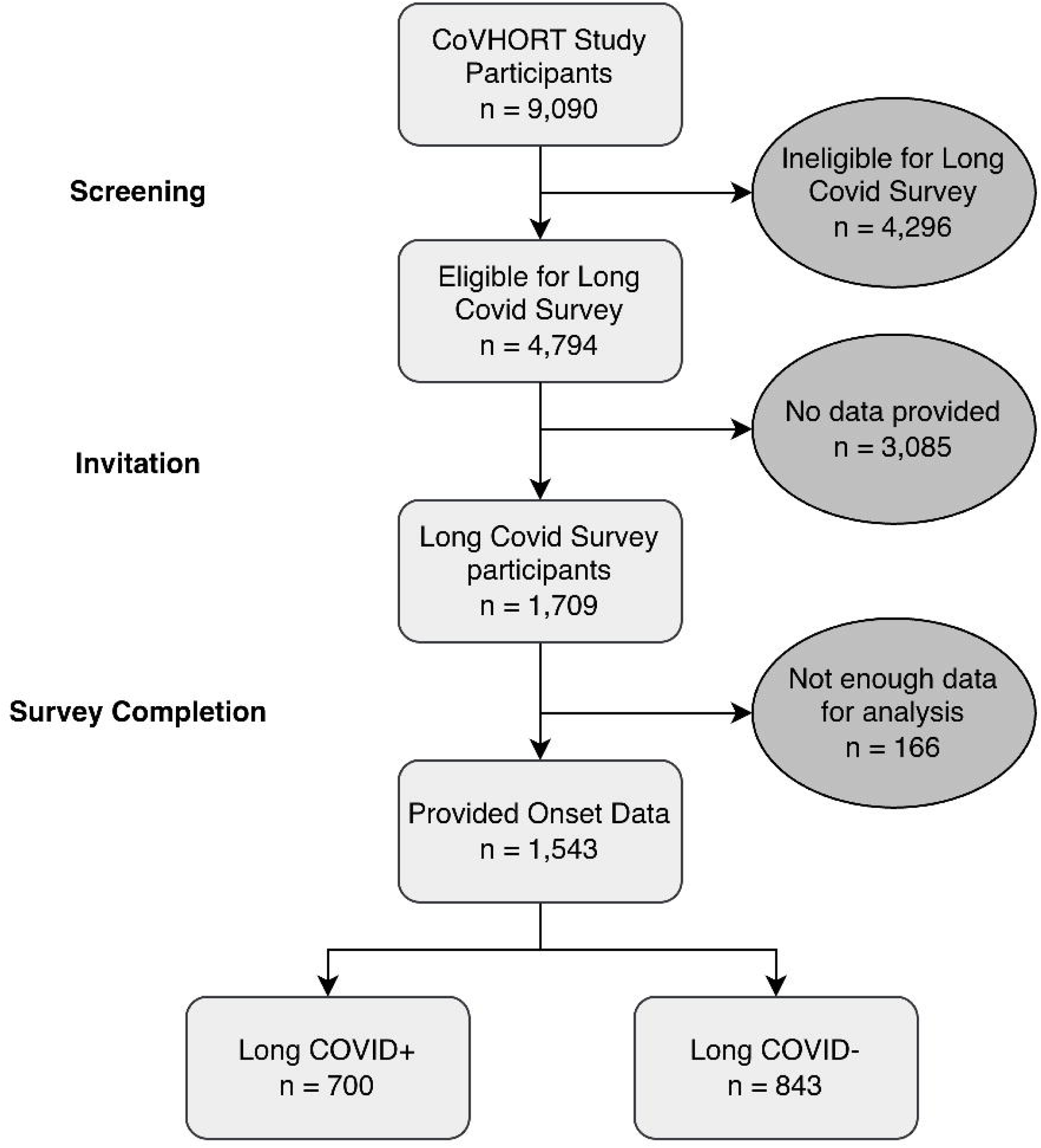

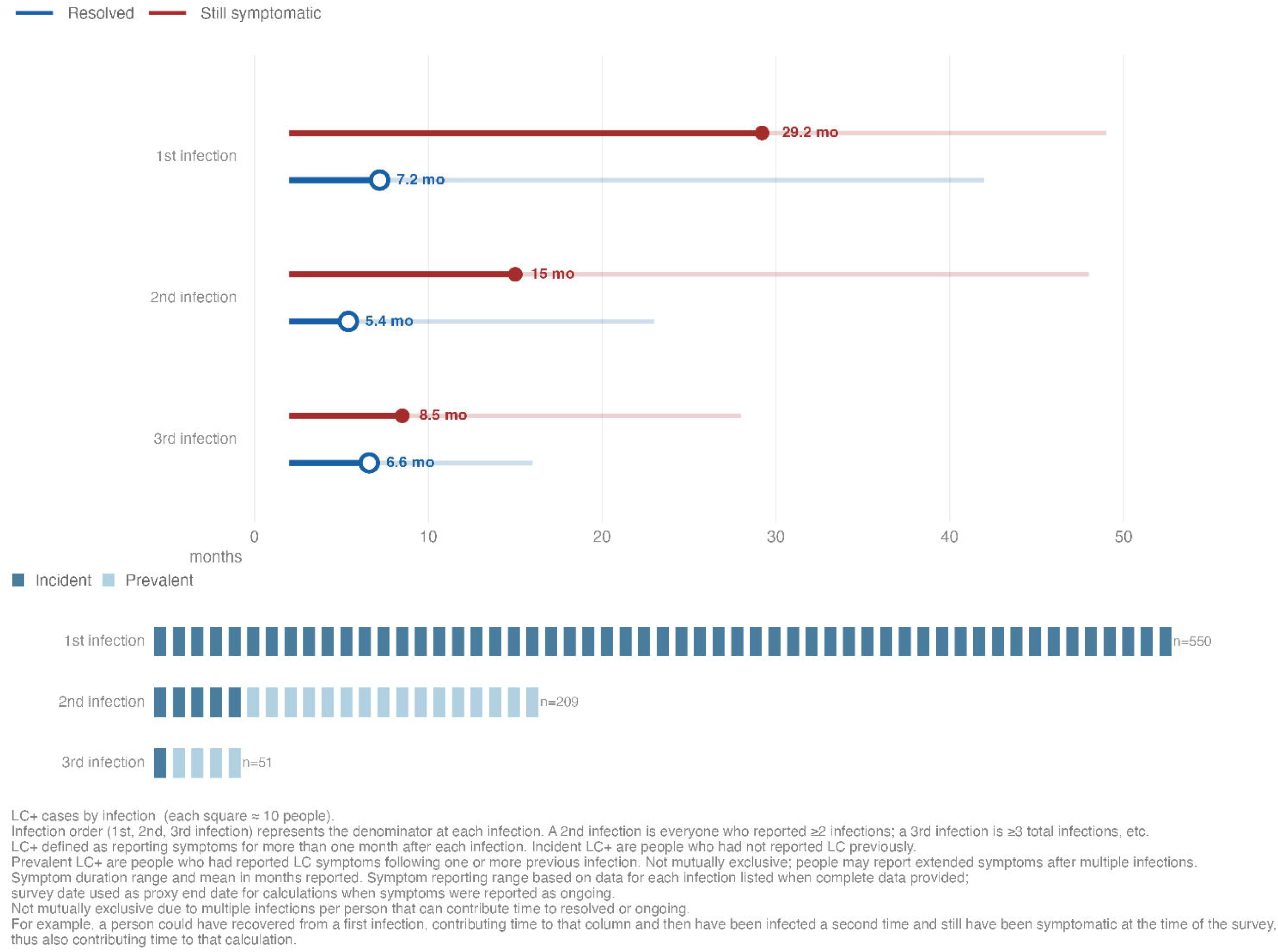

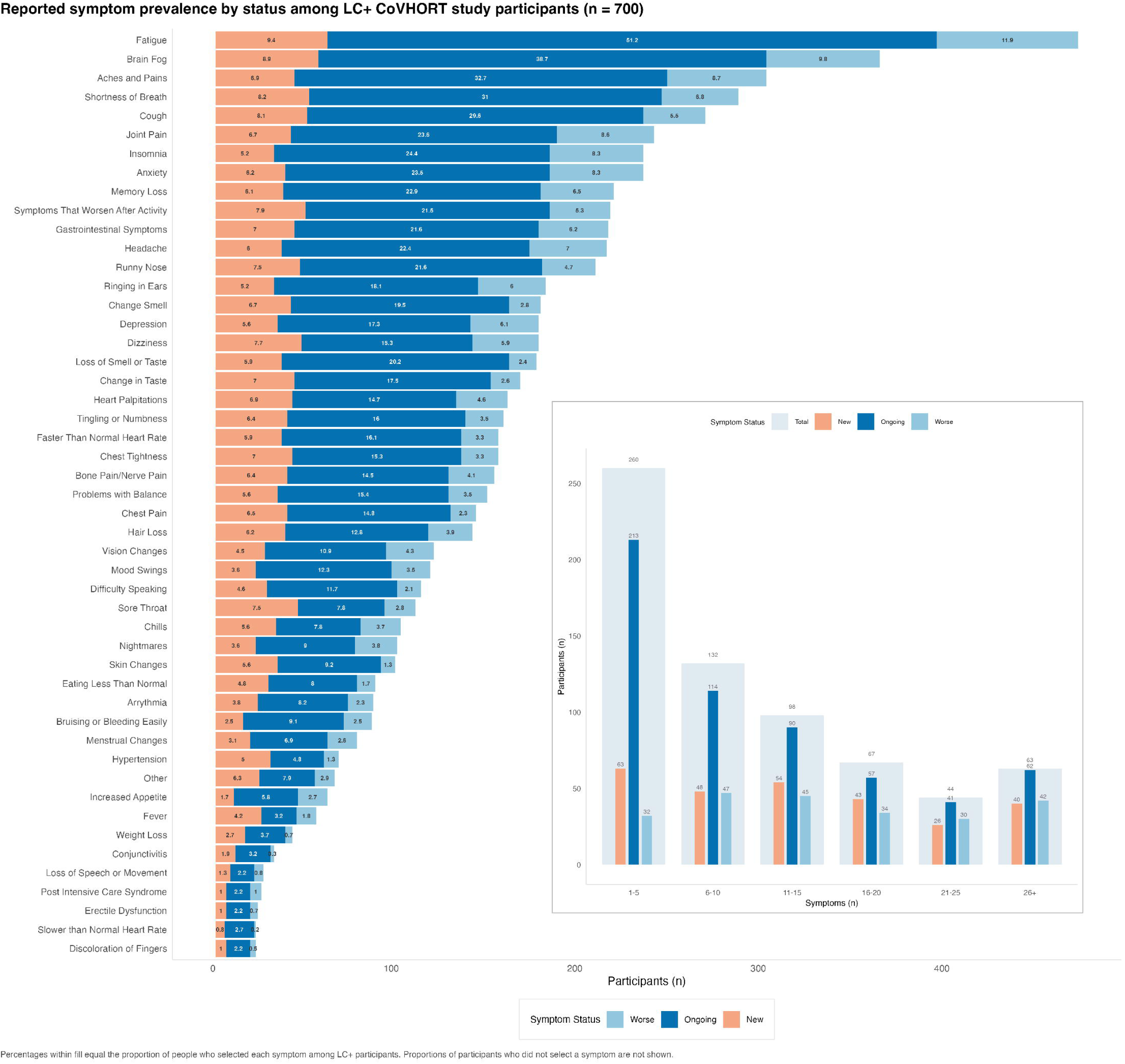

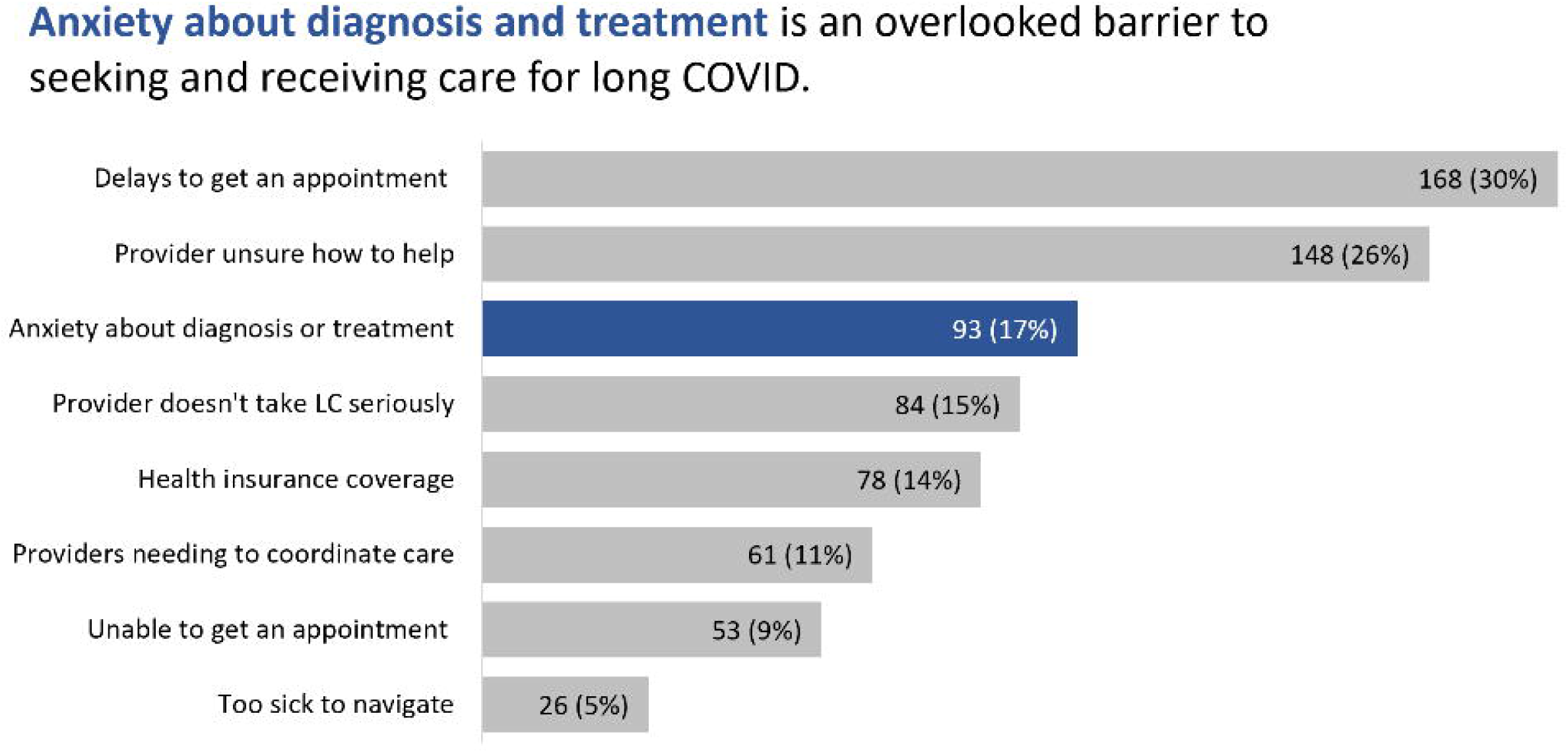

**Supplementary Table 1.**
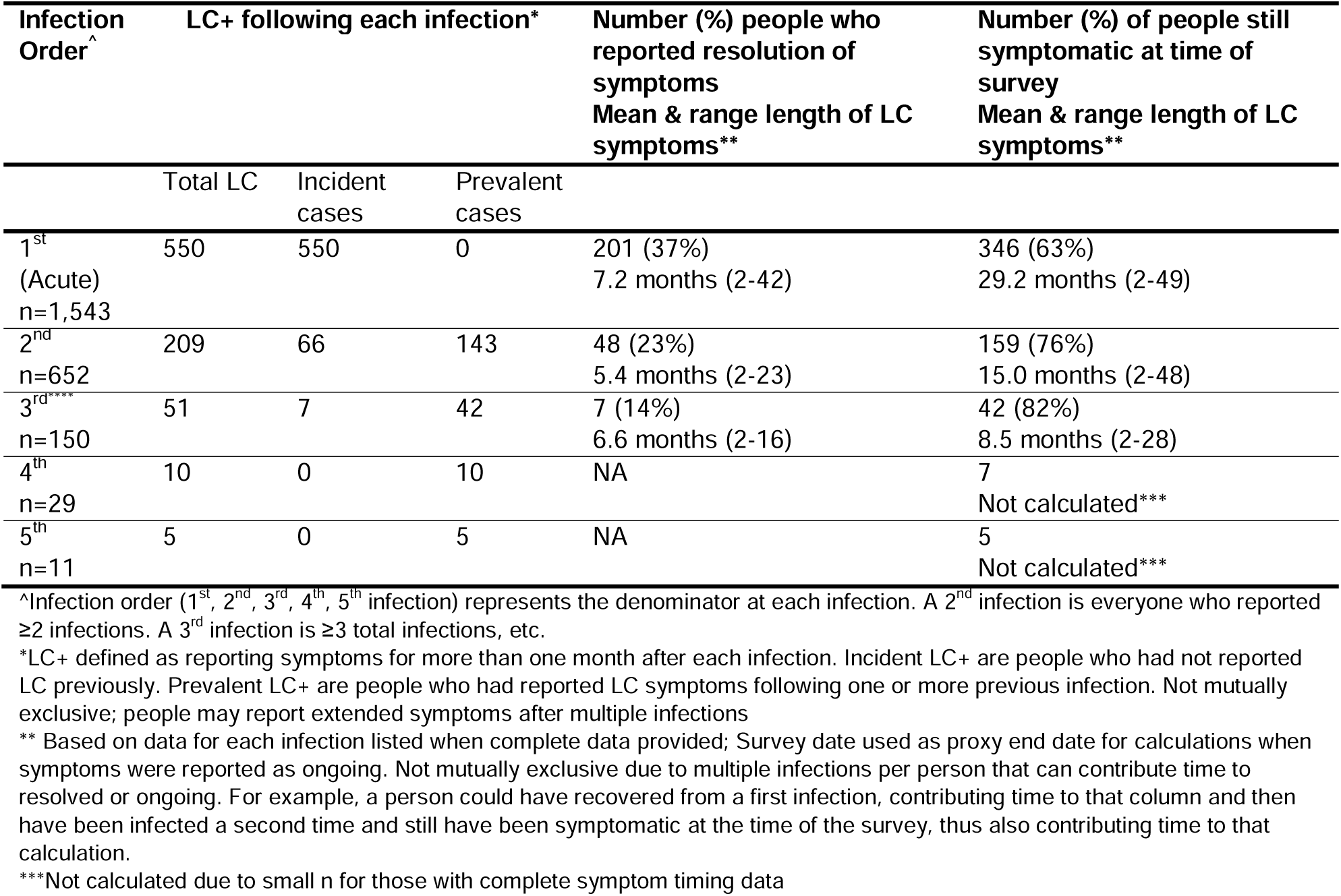
Length of symptoms and resolution among those with long COVID (n=700)

**Supplementary Table 2.**
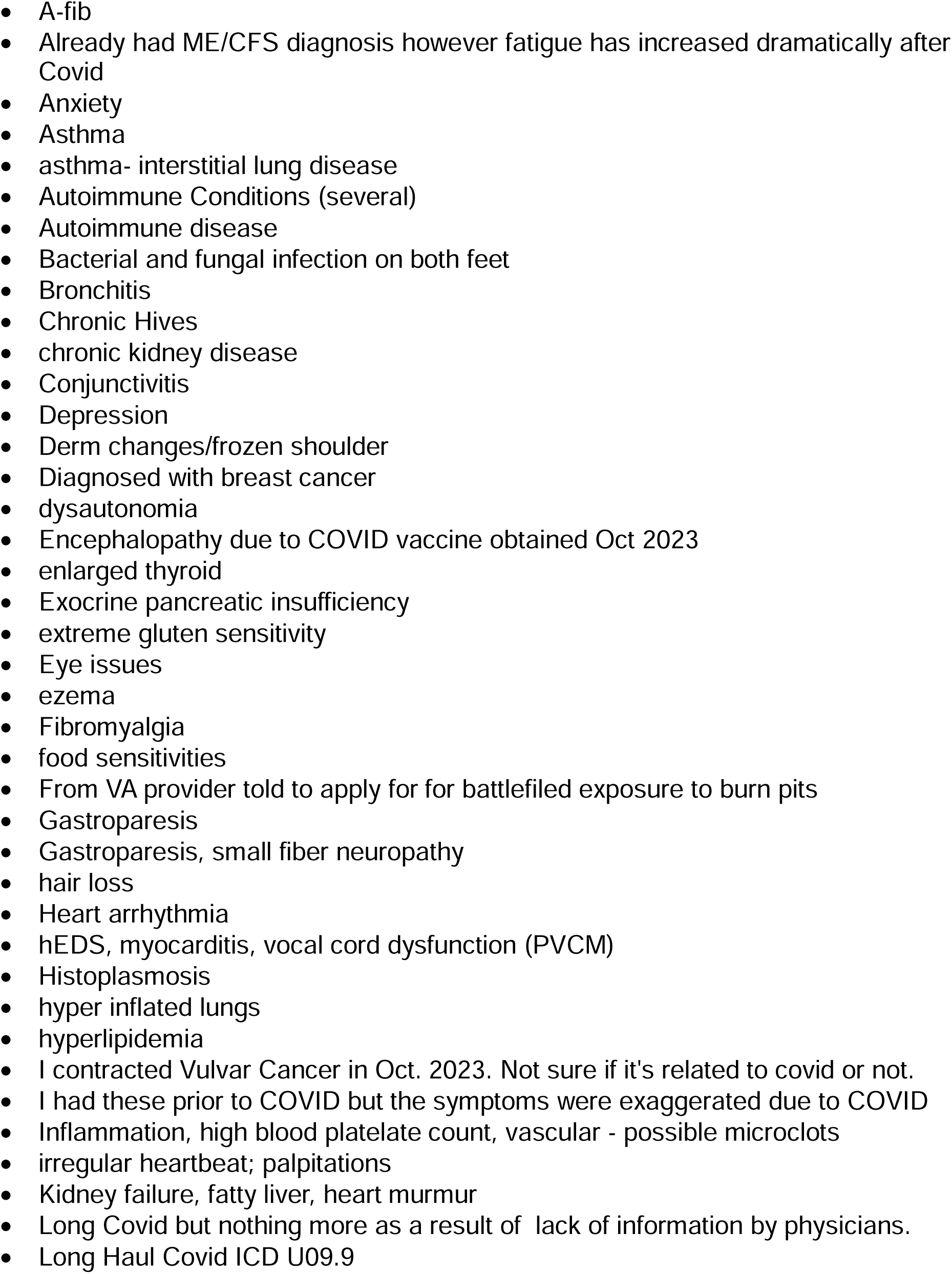
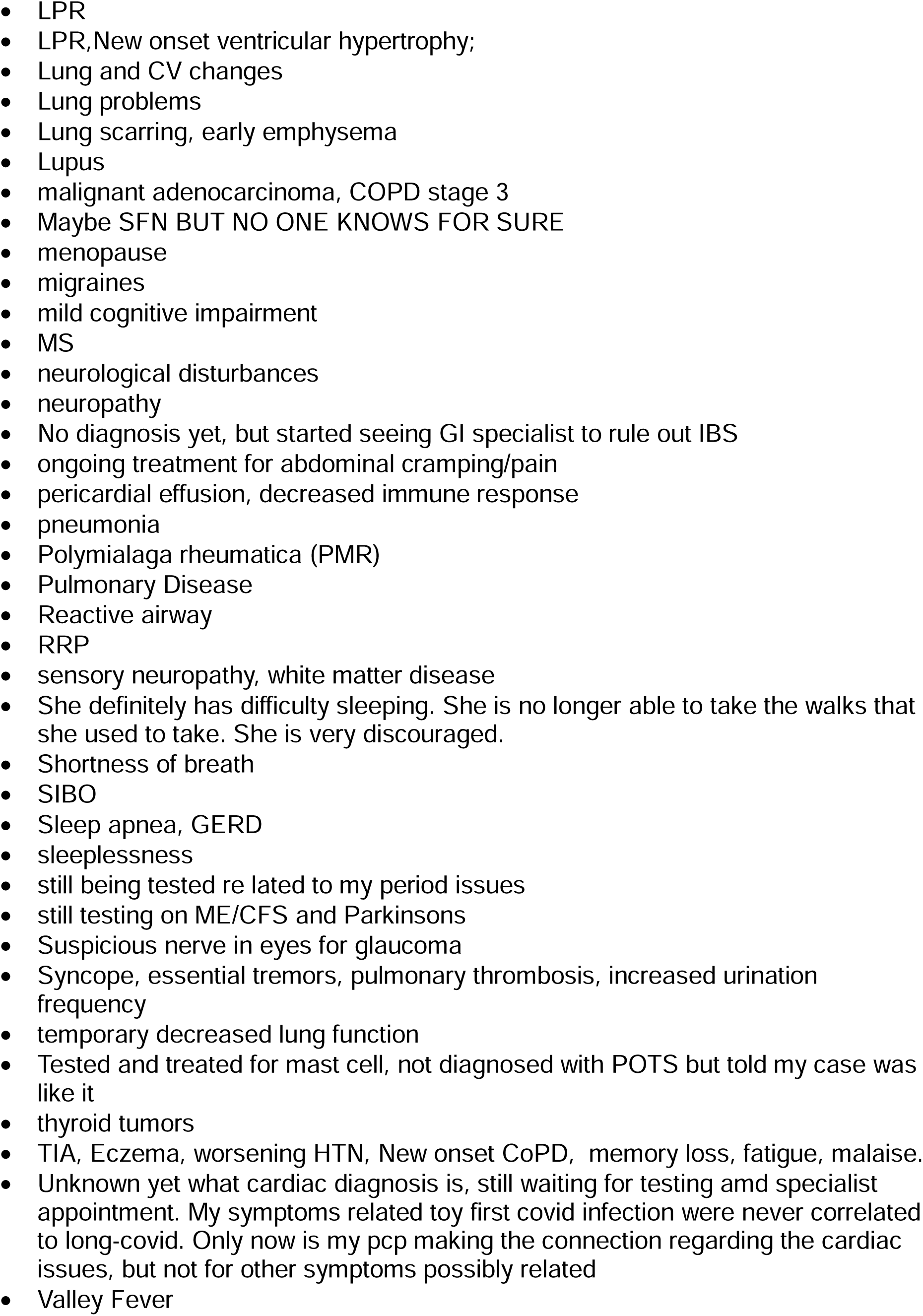
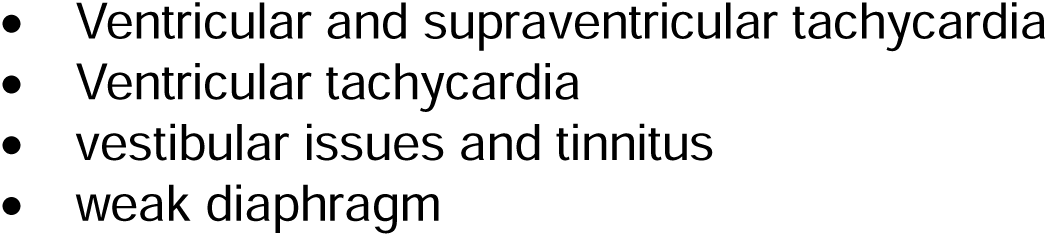
Other diagnoses provided by healthcare providers during LC visits.

